# Predicting Deprescribing of High-Risk Medications Using Provider EHR Use and Patient Characteristics

**DOI:** 10.64898/2026.07.25.26358746

**Authors:** Bowen Gu, Katharina Tabea Jungo, Julie C Lauffenburger, Niteesh K Choudhry, Thomas Isaac, John Zambrano, Jie Yang

## Abstract

Potentially inappropriate medications expose older adults to preventable harm, yet deprescribing remains difficult to implement consistently. Although electronic health record (EHR) interventions can reduce prescribing, health systems lack clear evidence about which routinely captured patient, primary care provider (PCP), and intervention-design factors predict medication discontinuation or dose tapering. Understanding these determinants is essential for targeting and scaling deprescribing support. In this study, we conducted the first machine-learning analysis of these trial data. We analyzed 2,979 adults aged 65 years or older and 158 structured EHR features spanning patient characteristics, PCP characteristics and EHR-use behaviors, and deprescribing-tool design. We compared eight models for predicting medication discontinuation or dose tapering and used SHAP to examine feature importance. TabPFN achieved the highest positive predictive value (67.71%), AUROC (74.30%), and AUPRC (60.95%), although overall predictability was moderate. Our findings show that even rich structured EHR data only moderately predict deprescribing, suggesting that important clinical determinants are not captured in routine fields. PCP EHR-use measures accounted for 19 of the 25 highest-ranked TabPFN features, although they also constituted most candidate predictors. The study provides the health system with an informative reference for predicting high-risk medication deprescribing. Future models should incorporate richer clinical context and undergo external validation before informing personalized deprescribing support.

## Introduction

Many older adults receive potentially-inappropriate medications for which potential harms outweigh benefits, and deprescribing (stopping or reducing dosage) is recommended.^1^ Prior studies have shown that deprescribing tools integrated in electronic health record (EHR) systems can reduce prescribing.^2,3^ These findings extend prior work identifying distinct EHR-use phenotypes among PCPs by suggesting that routine EHR-use behaviors vary among PCPs^4^ and may also be associated with responsiveness to deprescribing interventions.

Understanding how patient-, provider-, and deprescribing tool design-level factors captured in EHR data affect deprescribing is critical for scaling and targeting them. Thus, we used 8 machine learning methods to predict high-risk medication deprescribing and identify key predictors using EHR data.

## Methods

### Setting

We used data from a 16-arm adaptive cluster-randomized NUDGE-EHR trial.^2^ Eligible participants were PCPs and their patients aged ≥65 years prescribed either ≥90 pills of benzodiazepines or non-benzodiazepine sedative hypnotics or ≥90 pills of ≥2 strongly anticholinergic medications in the prior 180 days. The intervention consisted of enhanced EHR deprescribing alerts with order sets, patient handouts, and tapering algorithms, triggered either when ordering medication or opening an encounter. Across arms, alerts incorporated behavioral science features (follow-up boosters, priming messages, simplified language, precommitment, additional sign-off timing, alternative risk framing), while comparator arms received a standard alert or usual care.

### Dataset

The dataset includes PCP sociodemographic characteristics, EHR use variables (calculated once as the average over the 3 months before the PCP’s first index date and once over the 3 months before each patient’s index visit), patient sociodemographic and clinical characteristics, information on behavioral features included in EHR tools, and a composite deprescribing outcome, the trial’s primary outcome, defined as medication discontinuation or dose taper (**Appendix 1**). We used structured EHR data to build a scalable model from readily extractable features.

### Preprocessing

The final processed data contained 2,979 patients with 158 features (97.3%, 2,979/3,063 trial participants; 158/179 features) (**Figure S1, Appendix 2)**. All continuous features were normalized using standard scaling. Patients were randomly split into an 80% training set and a 20% test set.

### Model prediction

We utilized 8 advanced and most representative machine models for the deprescribing prediction. Specifically, we trained four regression models (logistic, lasso, ridge, and elastic net), two support vector machine (SVM) models (linear and radial basis function [RBF]), one boosting model (XGBoost),^5^ and one transformer-based AI foundation model (TabPFN).^6^ A random guessing baseline model that randomly categorized each test case as positive or negative with equal probabilities was tested for additional comparison. We also examined the explainability of the TabPFN model by using SHapley Additive exPlanations (SHAP), which assigned each feature an importance value for a particular prediction.^7^ We plotted the mean of the absolute SHAP values of the top features that contribute the most to the model’s prediction. Each model’s detailed parameter settings and package versions are available in **Appendix 3**. Model performance was evaluated using accuracy, precision (PPV), F1 score, recall, negative predictive value (NPV), AUROC, and AUPRC (**Appendix 2**). TabPFN was evaluated without preprocessing, as it requires none.^8^

## Results

Overall, the provider-, and deprescribing tool design-level factors included in our analysis did not strongly predict deprescribing decisions. TabPFN showed the most balanced performance with the highest PPV (67.71%), AUROC (74.30%) and AUPRC (60.95%), with accuracy and NPV among the highest. All models outperformed the baseline in accuracy, AUROC, and AUPRC, but most models had lower recall than the baseline (**Table 1)**. The logistic regression achieved the highest accuracy of 71.98%, together with the highest NPV of 73.44%. Linear SVM achieved the highest F1 of 49.42% and the highest recall of 62.56%. The RBF SVM predicted all test set samples as negative, indicating that the fitting was not ideal.

**Table 1.** Performance of all models and the baseline. The highest value of each column is highlighted in **bold**. The definitions of each metric are shown in **Appendix 2**.

| Model | Accuracy (%) | Precision (PPV) (%) | F1 (%) | Recall (%) | NPV (%) | AUROC (%) | AUPRC (%) |
| --- | --- | --- | --- | --- | --- | --- | --- |
| *Baseline (Random Guessing)* | 52.85 | 35.66 | 40.84 | 47.78 | 67.28 | 50.00 | 34.06 |
| *Logistic Regression* | **71.98** | 65.79 | 47.32 | 36.95 | **73.44** | 70.33 | 56.12 |
| *Lasso Logistic Regression* | 65.94 | 50.00 | 6.45 | 3.45 | 66.32 | 60.62 | 44.54 |
| *Ridge Logistic Regression* | 71.64 | 66.04 | 45.31 | 34.48 | 72.86 | 70.66 | 56.23 |
| *Elastic Net Logistic Regression* | 65.94 | 50.00 | 6.45 | 3.45 | 66.32 | 60.64 | 44.52 |
| *Linear SVM* | 56.38 | 40.84 | **49.42** | **62.56** | 73.33 | 57.79 | 42.89 |
| *RBF SVM* | 65.94 | 0.00 | 0.00 | 0.00 | 65.94 | 55.06 | 39.44 |
| *XGBoost* | 68.62 | 55.48 | 46.42 | 39.90 | 72.89 | 67.96 | 53.22 |
| *TabPFN* | 71.64 | **67.71** | 43.48 | 32.02 | 72.40 | **74.30** | **60.95** |

Figure 1 shows the SHAP values of the 25 most important features of TabPFN. 76.00% (19/25) of the top-rated features are related to PCP EHR use, 16.00% (4/25) are patient characteristics, and 8.00% (2/25) are PCP characteristics. This distribution is similar to the one among the 158 input features (67.09% (106/158) are PCP EHR use features, 15.19% (24/158) are patient characteristics, and 7.59% (12/158) are PCP characteristics).

**Figure 1.**
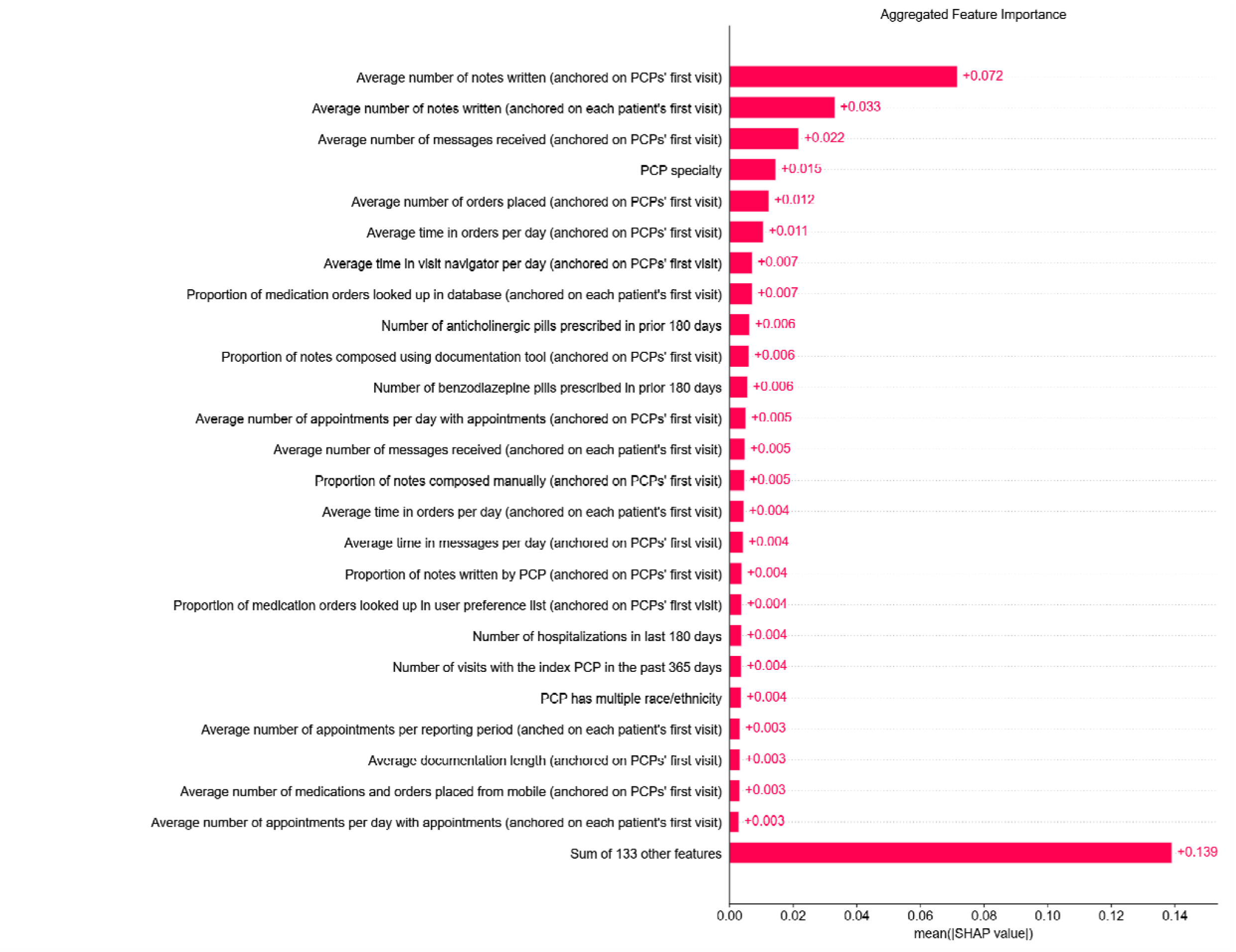
Mean absolute SHAP value of the input features on the TabPFN model.

## Discussion

This study evaluated eight models to predict deprescribing of high-risk medication using comprehensive patient and PCP features and is the first to apply machine learning on this trial data. Limitations include the use of a secondary analysis of a single trial dataset, a relatively small sample size, potential provider-level clustering, the absence of unstructured data, and a lack of external validation.

Despite rich patient, provider, and intervention data from a large trial, deprescribing decisions were only moderately predictable from structured EHR data, suggesting that important determinants of deprescribing may not be captured in routine EHR fields. Among all models, the transformer-based TabPFN model had the most stable performance. Nevertheless, provider EHR-use behaviors appeared more informative than patient characteristics or intervention design features. Although EHR-use variables dominated the highest-ranking features, they also represented most available predictors.

## Data Availability

All data produced in the present study are available upon reasonable request to the authors

## Acknowledgments

Research reported in this publication was supported by the National Institute on Aging of the National Institutes of Health under Award Number R33AG057388 to BWH (Choudhry/Lauffenburger PI). The content is solely the responsibility of the authors and does not necessarily represent the official views of the National Institutes of Health. JY was funded by PCORI ME-2022C1-25646, Goldberg Scholarship and Brigham Research Institute, National Institute on Aging RF1AG090405, and National Library of Medicine R01LM014667. KTJ was supported by a Postdoc.Mobility Fellowship from the Swiss National Science Foundation [P500PM_206728].

