## Supporting Material for "Predicting Deprescribing of High-Risk Medications Using Provider EHR Use and Patient Characteristics"

**Title**

Julie C Lauffenburger, PharmD, PhD,^1,2^

Niteesh K Choudhry, MD, PhD,^1,2^

Thomas Isaac, MD, MBA,^4^

John Zambrano, MD,^4^

Jie Yang, PhD, FACMI, FAMIA^1,2,5,6^

**Affiliations**

^1^ Division of Pharmacoepidemiology and Pharmacoeconomics, Department of Medicine, Brigham and Women’s Hospital, Boston, MA, USA

^2^ Harvard Medical School, Boston, MA, USA

^3^ Institute of Primary Healthcare, University of Bern, Bern, CH

^4^ Atrius Health, Newton, Massachusetts, USA

^5^ Harvard Data Science Initiative, Harvard University, Cambridge, MA, USA

^6^ Broad Institute of MIT and Harvard, Cambridge, MA, USA

### Appendix 1. Variables included in the analysis.

| **Feature ID** | **Feature Name** | **Description** |
| --- | --- | --- |
| **Patient Characteristics** | | |
| 1 | pat_age | Patient age |
| 2 | pat_sex | Patient sex |
| 3 | pat_ethnicity_White_yes | Patient race/ethnicity is White |
| 4 | pat_ethnicity_Hispanic or Latino_yes | Patient race/ethnicity is Hispanic or Latino |
| 5 | pat_ethnicity_Black or African American_yes | Patient race/ethnicity is Black or African American |
| 6 | pat_ethnicity_Patient Declined_yes | Patient declined race/ethnicity |
| 7 | pat_ethnicity_Other_yes | Patient race/ethnicity is Other |
| 8 | pat_ethnicity_Asian_yes | Patient race/ethnicity is Asian |
| 9 | pat_ethnicity_American Indian or Alaska Native_yes | Patient race/ethnicity is American Indian or Alaska Native |
| 10 | benzod_quantity | Number of benzodiazepine pills prescribed in prior 180 days |
| 11 | sedative_quantity | Number of non-sedative hypnotic pills prescribed in prior 180 days |
| 12 | anticholinergics_quantity | Number of anticholinergic pills prescribed in prior 180 days |
| 13 | Multi_classes | Number of high-risk medication classes used |
| 14 | index_Encounter_Type | Type of index visit with PCP |
| 15 | mild_cognitive_impairment | Mild cognitive impairment diagnosis |
| 16 | dementia | Dementia diagnosis |
| 17 | depression | Depression diagnosis |
| 18 | anxiety | Anxiety diagnosis |
| 19 | insomnia | Insomnia diagnosis |
| 20 | pat_chronic_pain | Chronic pain diagnosis |
| 21 | pat_PCP_visits_365 | Number of visits with the index PCP in the past 365 days |
| 22 | pat_last_base_visit_is_PCP_visit | Last visit with index PCP |
| 23 | pat_hosp_in_180_before_index | Number of hospitalizations in last 180 days |
| 24 | pat_ER_visits_in_180_before_inde | Number of ER visits in last 180 days |
| **Provider (PCP) Characteristics** | | |
| 25 | provider_type_Physician_yes | PCP type is physician |
| 26 | provider_type_Physician Assistant_yes | PCP type is physician assistant |
| 27 | provider_type_Osteopath_yes | PCP type is osteopath |
| 28 | provider_type_Nurse Practitioner_yes | PCP type is nurse practitioner |
| 29 | speciality | PCP specialty |
| 30 | provider_gender | PCP Sex |
| 31 | provider_ethnicity_White_yes | PCP race/ethnicity is White |
| 32 | provider_ethnicity_Asian_yes | PCP race/ethnicity is Asian |
| 33 | provider_ethnicity_Hispa_yes | PCP race/ethnicity is Hispanic |
| 34 | provider_ethnicity_Black_yes | PCP race/ethnicity is Black |
| 35 | provider_ethnicity_Multi_yes | PCP has multiple race/ethnicity |
| 36 | years_at_atrius | Number of years in health system |
| **PCP EHR Use (averages over last 3 reporting periods)** | | |
| 37 | pcp_panel_patient_count_avg3p | Average panel size (measured in the last 3 reporting periods before the index date) |
| 38 | dayswithappts_ratio_avg3p | Average number of days with appointments (anchored on PCPs' first visit) |
| 39 | apptsperday_ratio_avg3p | Average number of appointments per day with appointments (anchored on PCPs' first visit) |
| 40 | apptsperperiod_avg3p | Number of appointments per reporting period (anchored on PCPs' first visit) |
| 41 | apptsclosedperday_avg3p | Percentage of appointments closed on the same day (anchored on PCPs' first visit) |
| 42 | lengthdocappt_avg_avg3p | Average length of documentation per appointment (anchored on PCPs' first visit) |
| 43 | medsapptothers_ratio_avg3p | Average medications signed per appointment to others (anchored on PCPs' first visit) |
| 44 | medsapptself_ratio_avg3p | Average medications signed per appointment to self (anchored on PCPs' first visit) |
| 45 | ordersteamcontrib_avg3p | Average orders placed with team contributions (anchored on PCPs' first visit) |
| 46 | percentorder_smartsets_avg3p | Percent of orders from preference lists & ordersets (anchored on PCPs' first visit) |
| 47 | timeclinrevappt_avg3p | Average time in clinical review per appointment (anchored on PCPs' first visit) |
| 48 | timeinbasketappt_avg3p | Average time in in-basket per appointment (anchored on PCPs' first visit) |
| 49 | timenotesappt_avg3p | Average time in notes per appointment (anchored on PCPs' first visit) |
| 50 | timeordersappt_avg3p | Average time in orders per appointment (anchored on PCPs' first visit) |
| 51 | EHRusedaysappts_avg3p | Average time in EHR system per day with appointments (anchored on PCPs' first visit) |
| 52 | EHRpjtimedaysappts_avg3p | Average time in EHR system outside of 7a-7p per day with appointments (anchored on PCPs' first visit) |
| 53 | EHRunschdaysappts_avg3p | Average time in EHR per unscheduled day (anchored on PCPs' first visit) |
| 54 | avgspeedbutton_avg3p | Average number of diagnosis speed buttons (anchored on PCPs' first visit) |
| 55 | doculength_avg3p | Average documentation length (anchored on PCPs' first visit) |
| 56 | poolmessages_avg3p | Proportion of messages completed (anchored on PCPs' first visit) |
| 57 | lookupsource_db_avg3p | Proportion of medication orders looked up in database (anchored on PCPs' first visit) |
| 58 | lookupsource_systlist_avg3p | Proportion of medication orders looked up in system preference list (anchored on PCPs' first visit) |
| 59 | lookupsource_userlist_avg3p | Proportion of medication orders looked up in user preference list (anchored on PCPs' first visit) |
| 60 | medsappt_avg3p | Average number of medications signed per appointment (anchored on PCPs' first visit) |
| 61 | placedonmobile_avg3p | Average number of medications and orders placed from mobile (anchored on PCPs' first visit) |
| 62 | composed_copypaste_avg3p | Proportion of notes composed using Copy/Paste (anchored on PCPs' first visit) |
| 63 | composed_manual_avg3p | Proportion of notes composed manually (anchored on PCPs' first visit) |
| 64 | composed_notewriter_avg3p | Proportion of notes composed using NoteWriter (anchored on PCPs' first visit) |
| 65 | composed_smarttool_avg3p | Proportion of notes composed using documentation tool (anchored on PCPs' first visit) |
| 66 | composed_transcription_avg3p | Proportion of notes composed using Transcription (anchored on PCPs' first visit) |
| 67 | composed_voicerec_avg3p | Proportion of notes composed using VoiceRecognition (anchored on PCPs' first visit) |
| 68 | contr_notes_other_avg3p | Proportion of notes written by others (anchored on PCPs' first visit) |
| 69 | contr_notes_pcp_avg3p | Proportion of notes written by PCP (anchored on PCPs' first visit) |
| 70 | avgnotespeedbutton_avg3p | Average number of note speed buttons (anchored on PCPs' first visit) |
| 71 | avgibmessagesreceived_avg3p | Average number of messages received (anchored on PCPs' first visit) |
| 72 | numnoteswritten_avg3p | Average number of notes written (anchored on PCPs' first visit) |
| 73 | numordersplaced_avg3p | Average number of orders placed (anchored on PCPs' first visit) |
| 74 | dbordersourcelookup_avg3p | Average number of other order preference list/database lookup source – database (anchored on PCPs' first visit) |
| 75 | systemordersourcelookup_avg3p | Average number of other order preference list/database lookup source – system preference list (anchored on PCPs' first visit) |
| 76 | userordersourcelookup_avg3p | Average number of other order preference list/database lookup source – user preference list (anchored on PCPs' first visit) |
| 77 | progressnotelength_avg3p | Average progress note length (anchored on PCPs' first visit) |
| 78 | quickactionsavailable_avg3p | Average number of QuickActions available (anchored on PCPs' first visit) |
| 79 | quickactionsusedperday_avg3p | Average number of QuickActions used per day (anchored on PCPs' first visit) |
| 80 | smartphrasescreated_avg3p | Average number of SmartPhrases created by PCP (anchored on PCPs' first visit) |
| 81 | timeclinicalreview_day_avg3p | Average time in clinical review per day (anchored on PCPs' first visit) |
| 82 | timeinbasket_day_avg3p | Average time in messages per day (anchored on PCPs' first visit) |
| 83 | timenotes_day_avg3p | Average time in notes per day (anchored on PCPs' first visit) |
| 84 | timeorders_day_avg3p | Average time in orders per day (anchored on PCPs' first visit) |
| 85 | timevisitnavigator_day_avg3p | Average time in visit navigator per day (anchored on PCPs' first visit) |
| 86 | timeoutsidehours_avg3p | Average time outside scheduled hours (anchored on PCPs' first visit) |
| 87 | tatpatientadvice_min_avg3p | Average turnaround time – patient medical advice request (in minutes) (anchored on PCPs' first visit) |
| 88 | tatrxauth_min_avg3p | Average turnaround time – Rx auth (in minutes) (anchored on PCPs' first visit) |
| 89 | unchangeddefaults_avg3p | Proportion of unchanged defaults from preference list (anchored on PCPs' first visit) |
| **EHR Tool Features** | | |
| 90 | tool_num_collapsed_orderentry_v2 | Order entry alert (yes/no) - EHR deprescribing tool |
| 91 | tool_num_collapsed_openen_v2 | Open encounter alert (yes/no) - EHR deprescribing tool |
| 92 | tool_num_collapsed_booster_v2 | Follow-up booster (yes/no) - EHR deprescribing tool |
| 93 | tool_num_collapsed_coldstate_v2 | Cold-state priming (yes/no) - EHR deprescribing tool |
| 94 | tool_num_collapsed_simplified_v2 | Simplification (yes/no) - EHR deprescribing tool |
| 95 | tool_num_collapsed_signoff_v2 | Additional sign-off (yes/no) - EHR deprescribing tool |
| 96 | tool_num_collapsed_precommit_v2 | Pre-commitment (yes/no) - EHR deprescribing tool |
| 97 | tool_num_collapsed_riskfr_v2 | Alternative risk framing (yes/no) - EHR deprescribing tool |
| **Visit Timing** | | |
| 98 | is_sunday | Index visit was on a Sunday |
| 99 | is_monday | Index visit was on a Monday |
| 100 | is_tuesday | Index visit was on a Tuesday |
| 101 | is_wednesday | Index visit was on Wednesday |
| 102 | is_thursday | Index visit was on a Thursday |
| 103 | is_friday | Index visit was on a Friday |
| 104 | is_saturday | Index visit was on a Saturday |
| **PCP Panel / Exposure** | | |
| 105 | number_eligible_pat | Number of eligible patients per PCP |
| **PCP EHR Use (same definitions as avg3p but anchored to PCP cluster’s first patient** | | |
| 106 | pcp_panel_patient_count_avgfirst | Average panel size (measured as an average of the last 3 reporting periods before the index date of the first patient per PCP cluster) (Patient-level) |
| 107 | dayswithappts_ratio_avgfirst | Average number of days with appointments (anchored on each patient's first visit) |
| 108 | apptsperday_ratio_avgfirst | Average number of appointments per day with appointments (anchored on each patient's first visit) |
| 109 | apptsperperiod_avgfirst | Average number of appointments per reporting period (anched on each patient's first visit) |
| 110 | apptsclosedperday_avgfirst | Percentage of appointments closed on the same day (anchored on each patient's first visit) |
| 111 | lengthdocappt_avg_avgfirst | Average length of documentation per appointment (anchored on each patient's first visit) |
| 112 | medsapptothers_ratio_avgfirst | Average medications signed per appointment to others (anchored on each patient's first visit) |
| 113 | medsapptself_ratio_avgfirst | Average medications signed per appointment to self (anchored on each patient's first visit) |
| 114 | ordersteamcontrib_avgfirst | Average orders placed with team contributions (anchored on each patient's first visit) |
| 115 | percentorder_smartsets_avgfirst | Percent of orders from preference lists and ordersets (anchored on each patient's first visit) |
| 116 | timeclinrevappt_avgfirst | Average time in clinical review per appointment (anchored on each patient's first visit) |
| 117 | timeinbasketappt_avgfirst | Average time in messages per appointment (anchored on each patient's first visit) |
| 118 | timenotesappt_avgfirst | Average time in notes per appointment (anchored on each patient's first visit) |
| 119 | timeordersappt_avgfirst | Average time in orders per appointment (anchored on each patient's first visit) |
| 120 | EHRusedaysappts_avgfirst | Average time in EHR system per day with appointments (anchored on each patient's first visit) |
| 121 | EHRpjtimedaysappts_avgfirst | Average time in EHR system outside of 7a–7p per day with appointments (anchored on each patient's first visit) |
| 122 | EHRunschdaysappts_avgfirst | Average time in EHR per unscheduled day (anchored on each patient's first visit) |
| 123 | avgspeedbutton_avgfirst | Average number of diagnosis speed buttons (anchored on each patient's first visit) |
| 124 | doculength_avgfirst | Average documentation length (anchored on each patient's first visit) |
| 125 | poolmessages_avgfirst | Proportion of messages completed (anchored on each patient's first visit) |
| 126 | lookupsource_db_avgfirst | Proportion of medication orders looked up in database (anchored on each patient's first visit) |
| 127 | lookupsource_systlist_avgfirst | Proportion of medication orders looked up in system preference list (anchored on each patient's first visit) |
| 128 | lookupsource_userlist_avgfirst | Proportion of medication orders looked up in user preference list (anchored on each patient's first visit) |
| 129 | medsappt_avgfirst | Average number of medications signed per appointment (anchored on each patient's first visit) |
| 130 | placedonmobile_avgfirst | Average number of medications and orders placed from mobile (anchored on each patient's first visit) |
| 131 | composed_copypaste_avgfirst | Proportion of notes composed using Copy/Paste (anchored on each patient's first visit) |
| 132 | composed_manual_avgfirst | Proportion of notes composed manually (anchored on each patient's first visit) |
| 133 | composed_notewriter_avgfirst | Proportion of notes composed using NoteWriter (anchored on each patient's first visit) |
| 134 | composed_smarttool_avgfirst | Proportion of notes composed using documentation tool (anchored on each patient's first visit) |
| 135 | composed_transcription_avgfirst | Proportion of notes composed using Transcription (anchored on each patient's first visit) |
| 136 | composed_voicerec_avgfirst | Proportion of notes composed using VoiceRecognition (anchored on each patient's first visit) |
| 137 | contr_notes_other_avgfirst | Proportion of notes written by others (anchored on each patient's first visit) |
| 138 | contr_notes_pcp_avgfirst | Proportion of notes written by PCP (anchored on each patient's first visit) |
| 139 | avgnotespeedbutton_avgfirst | Average number of note speed buttons (anchored on each patient's first visit) |
| 140 | avgibmessagesreceived_avgfirst | Average number of messages received (anchored on each patient's first visit) |
| 141 | numnoteswritten_avgfirst | Average number of notes written (anchored on each patient's first visit) |
| 142 | numordersplaced_avgfirst | Average number of orders placed (anchored on each patient's first visit) |
| 143 | dbordersourcelookup_avgfirst | Average number of other order preference list/database lookup source – database (anchored on each patient's first visit) |
| 144 | systemordersourcelookup_avgfirst | Average number of other order preference list/database lookup source – system preference list (anchored on each patient's first visit) |
| 145 | userordersourcelookup_avgfirst | Average number of other order preference list/database lookup source – user preference list (anchored on each patient's first visit) |
| 146 | progressnotelength_avgfirst | Average progress note length (anchored on each patient's first visit) |
| 147 | quickactionsavailable_avgfirst | Average number of QuickActions available (anchored on each patient's first visit) |
| 148 | quickactionsusedperday_avgfirst | Average number of QuickActions used per day (anchored on each patient's first visit) |
| 149 | smartphrasescreated_avgfirst | Average number of SmartPhrases created by PCP (anchored on each patient's first visit) |
| 150 | timeclinicalreview_day_avgfirst | Average time in clinical review per day (anchored on each patient's first visit) |
| 151 | timeinbasket_day_avgfirst | Average time in messages per day (anchored on each patient's first visit) |
| 152 | timenotes_day_avgfirst | Average time in notes per day (anchored on each patient's first visit) |
| 153 | timeorders_day_avgfirst | Average time in orders per day (anchored on each patient's first visit) |
| 154 | timevisitnavigator_day_avgfirst | Average time in visit navigator per day (anchored on each patient's first visit) |
| 155 | timeoutsidehours_avgfirst | Average time outside scheduled hours (anchored on each patient's first visit) |
| 156 | tatpatientadvice_min_avgfirst | Average turnaround time – patient medical advice request (in minutes) (anchored on each patient's first visit) |
| 157 | tatrxauth_min_avgfirst | Average turnaround time – Rx auth (in minutes) (anchored on each patient's first visit) |
| 158 | unchangeddefaults_avgfirst | Proportion of unchanged defaults from preference list (anchored on each patient's first visit) |

### Appendix 2. Description of data preprocessing.

We first removed all features with data missingness ≥5% (n=21 features) and then removed patients whose rows still had missing data (n=84). After that, the remaining features underwent the following feature transformation process: all categorical features (e.g., ethnicity) were split into multiple subcategories using one-hot encoding. All binary features (e.g., gender) were coded into ones and zeros. All continuous features (e.g., age) were normalized using standard scaling. Patients were randomly split into an 80% training set and a 20% test set.

### Appendix 3. Definition of evaluation metrics

*Accuracy*: Proportion of all predictions that are correct. Measures overall classification performance.

*Precision (PPV)*: Among all cases predicted as positive, the proportion that is truly positive. Reflects the reliability of positive predictions.

*F1*: Harmonic mean of precision and recall. Balances false positives and false negatives in a single metric.

*Recall***:** Among all truly positive cases, the proportion correctly identified as positive. Measures the ability to detect positives.

*NPV (negative predictive value)*: Among all cases predicted as negative, the proportion that are truly negative. Reflects the reliability of negative predictions.

*AUROC (area under the receiver operating characteristic curve)*: Threshold-independent measure of a model's ability to distinguish positive from negative cases.

*AUPRC (area under the precision–recall curve)*: Threshold-independent measure summarizing the tradeoff between precision and recall across all classification thresholds.

Given **TP** = True Positives, **FP** = False Positives, **TN** = True Negatives, **FN** = False Negatives, the above metrics can be defined as follows:

| **Metric** | **Equation** |
| --- | --- |
| Precision (PPV) | $\frac{TP}{TP+FP}$ |
| F1 | $\frac{2TP}{2TP+FP+FN}$ |
| Recall | $\frac{TP}{TP+FN}$ |
| NPV | $\frac{TN}{TN+FN}$ |

### Appendix 4. Parameters used and package versions for model training

For all regression models, we set *max_iter* = 100,000 with *random_state* = 42. The vanilla logistic regression and the elastic net regression used the *lbfgs* solver, and the lasso regression and the ridge regression used the *sage* solver. For the elastic net regression, the *l1_ratio* is set to 0.5 (i.e., half L1 and half L2 regularization). For both SVM models, we set *max_iter* = 100,000 with *random_state* = 42. For the XGBoost model, we set *n_estimators* = 100, *max_depth* = 6, *random_state* = 42, and *eval_metric* = "auc". For the TabPFN model, we used TabPFN-2.6, and the TabPFN classifier was initialized using default settings. The packages used to initialize the regression models and the SVM models were *scikit-learn* (v 1.6.1). For the XGBoost model, *xgboost* (v 3.0.2) was used. For the TabPFN model, *tabpfn* (v 7.0.1) was used. For the SHAP feature importance plot, the tabpfn-extension package (v 0.3.0) was used.

### Figure S1. Data preprocessing flowchart

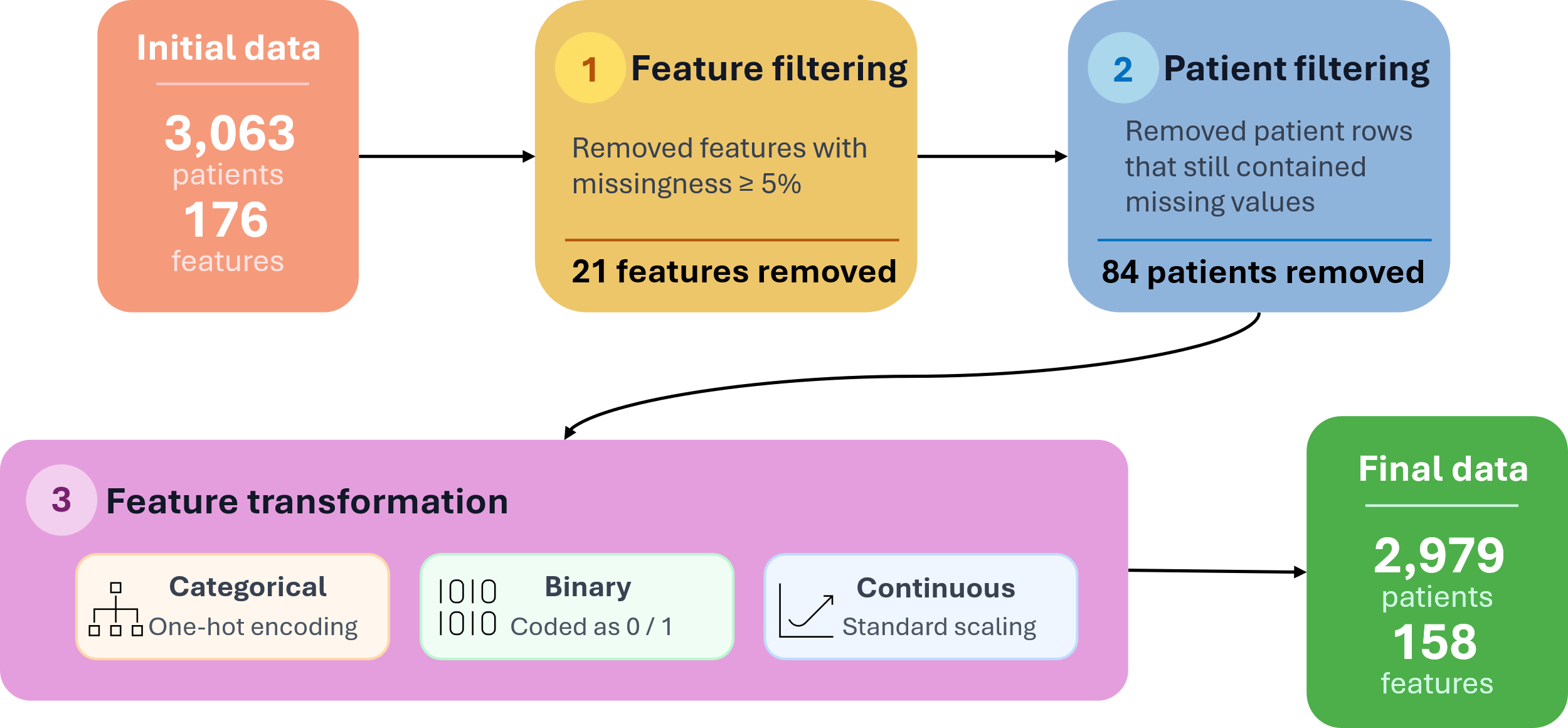
